# Can intersectionality help with understanding and tackling health inequalities? Perspectives of professional stakeholders

**DOI:** 10.1101/2020.10.26.20217463

**Authors:** Daniel Holman, Sarah Salway, Andrew Bell, Brian Beach, Adewale Adebajo, Nuzhat Ali, Jabeer Butt

## Abstract

The concept of ‘intersectionality’ is increasingly employed within public health arenas, particularly in North America, and is often heralded as offering great potential to advance health inequalities research and action. Given persistently poor progress towards tackling health inequalities, and recent calls to reframe this agenda in the UK and Europe, the possible contribution of intersectionality deserves attention. Yet, no existing research has examined professional stakeholder understandings and perspectives on applying intersectionality to this field. In this paper we seek to address that gap, drawing upon a consultation survey and workshop undertaken in the UK. The survey included both researchers (n=53) and practitioners (n=20) with varied roles and levels of engagement in research and evaluation. Topics included: familiarity with the term and concept ‘intersectionality’, relevance to health inequalities work, and issues shaping its uptake. Respondents were also asked to comment on two specific policy suggestions; targeting and tailoring interventions to intersectional sub-groups, and evaluating the intersectional effects of policies. 23 people attended the face-to-face workshop. The aims of the workshop were to: share examples of applying intersectionality within health inequalities research and practice; understand the views of research and practice colleagues on potential contributions and challenges; and identify potential ways to promote intersectional approaches. Findings indicated a generally positive response to the concept and a cautiously optimistic assessment that intersectional approaches could be valuable. However, opinions were mixed and various challenges were raised, especially around whether intersectionality research is necessarily critical and transformative and, accordingly, how it should be operationalised methodologically. Nonetheless, there was general agreement that intersectionality is concerned with diverse inequalities and the systems of power that shape them. We position intersectionality within the wider context of health inequalities policy and practice, suggesting potential ways forward for the approach in the UK context.

## Background

Kimberlé Crenshaw (1989) originally developed the idea of ‘intersectionality’ to highlight the ways in which prevailing legal and policy conceptions of discrimination overlooked the experiences of Black American women. Despite contestation on exactly what intersectionality is, there is a general agreement that it ‘references the critical insight that race, class, gender, sexuality, ethnicity, nation, ability, and age operate not as unitary, mutually exclusive entities, but rather as reciprocally constructing phenomena’ (Collins, 2015). Intersectionality is increasingly suggested as an innovative framework with the potential to advance understanding of, and action on, health inequalities, particularly by scholars in North America (Bauer, 2014; Bowleg, 2012; Hankivsky et al., 2017, 2014; Kapilashrami and Hankivsky, 2018). In particular, scholars have argued that intersectionality can illuminate diverse inequalities (Bowleg, 2012) and how power structures and processes give rise to them (Kapilashrami and Hankivsky, 2018). As a policy framework, Hankivksy et al. (2014) argues that intersectionality encourages critical reflection to move beyond singular categories, foregrounds issues of equity, and is innovative in highlighting processes of stigmatisation and the operation of power in policy making, offering various applied examples. Given persistently poor progress towards tackling health inequalities, and recent calls to reframe this agenda in the UK and Europe, the possible contribution of intersectionality deserves considered attention.

Regarded by many as a policy failure in England and beyond, (Bambra et al., 2011; Godding, 2014; Mackenbach, 2020, 2011; Marmot et al., 2020; Popay et al., 2010) two repeatedly highlighted limitations of health inequalities strategy are particularly pertinent. First is ‘life-style drift’; the tendency of policy initiatives to predominantly invest in individual behavioural interventions rather than address the ‘upstream’ social, political and economic determinants of poor health (Baum, 2011; Baum and Fisher, 2014; Whitehead, 2012). Intersectionality contrasts sharply with this approach by being fundamentally concerned with exposing and challenging deep-seated structures of discrimination. Second is the predominant focus on inequalities in health between groups defined by single axes of difference – most commonly socioeconomic measures – while failing to recognise other dimensions of identity and disadvantage (Ingleby, 2012; King et al., 2019; Salway et al., 2014).

Intersectionality takes as its starting point the recognition that social positions and identities are multiple and seeks to reveal the interconnected systems of subordination that together influence people’s life chances. There have been growing calls to find new ways of ‘framing’ health inequalities; to refresh current approaches to theorising and communicating their nature, causes and potential solutions within academic, public and policy arenas (Lundberg, 2020; Scambler, 2012; Smith and Schrecker, 2015).

Crenshaw drew on court cases, together with analysis of feminist and anti-racist theory and activism, to argue that dominant ways of describing and understanding discrimination are inadequate since they limit inquiry to the experiences of “otherwise-privileged members of the group” i.e. Black people who are men, or women who are White. This approach produces “a distorted analysis of racism and sexism because the operative conceptions of race and sex become grounded in experiences that actually represent only a subset of a much more complex phenomenon” (Crenshaw, 1989, p. 140). Crenshaw later elaborated how social class, age, sexuality and migrant status are other dimensions of identity where structures of power and discriminatory processes intersect (Crenshaw, 1991). Without a framework that acknowledges these intersectional experiences they are rendered invisible and their origins misunderstood. Further, policy and action that focuses on one social attribute at a time “limits remedial relief to minor adjustments within an established hierarchy” (Crenshaw, 1989, p. 145) and is a “trickle down approach to social justice” (Crenshaw, 2016) because it assumes that single attribute approaches are sufficiently inclusive.

Following Crenshaw, several researchers have argued for the utility of intersectionality within health inequalities work (Bauer, 2014; Bowleg, 2012; Hankivsky et al., 2014; Kapilashrami and Hankivsky, 2018; Lopez and Gadsden, 2016). Hankivsky et al. (2014) offer several examples from the Canadian context where the application of intersectionality has led to effective policy actions, such as in relation to HIV testing and prevention, aboriginal health and palliative care. Bowleg (2012) suggests that intersectionality can encourage us to examine the substantial heterogeneity within taken-for-granted categories, such as ‘women’, and the interplay of micro-level with macro-level factors producing disparate health outcomes (2012:1268). She summarises five main benefits of intersectionality for public health: it provides a unifying framework for scholars already interested in intersections of inequality; it acknowledges health inequalities as complex and multidimensional; its focus on macro-level factors is more likely to affect the fundamental causes of health inequalities; it informs the development of targeted and cost-effective interventions and policies, and; it supports the collection and analysis of rich socio-demographic and health data. Sen et al. (2009: 412) argue that:

> By giving precise insights into who is affected and how in different settings, [intersectionality] provides a scalpel for policies rather than the current hatchet. It enables policies and programmes to identify whom to focus on, whom to protect, what exactly to promote and why. It also provides a simple way to monitor and evaluate the impact of policies and programmes on different sub-groups from the most disadvantaged through the middle layers to those with particular advantages.

Yet such claims have so far not been properly explored with health inequalities researchers and those working in the policy and practice space, especially in the UK context. This is important because there is potentially a wide gap between the potential and rationale for intersectionality and the extent to which it is workable in practice. Nonetheless, ideas around intersectionality and health inequalities have now begun to emerge within high profile policy-facing work, with the recent Marmot et al. (2020) report of health equity in England recent report stating that “intersections between socioeconomic status, ethnicity and racism intensify inequalities in health for ethnic groups,” (p.23) and that “the cumulative experiences of multiple forms of disadvantage interact with and are exacerbated by features of the communities in which people live” (p.94).

Past work cautions against any simplistic expectation that a new concept will straightforwardly impact upon the ways in which health inequalities are understood or addressed. Health inequalities are recognised as a complex and ‘wicked’ problem; cutting across traditional organisational boundaries with diffuse responsibility and great scope for debate around what should be done and what counts as robust and relevant evidence (Blackman et al., 2006; Exworthy et al., 2006). Policy-making in such areas, far from being a technical exercise, is a process of dialogue, negotiation and ‘knowledge interaction’, with power relationships, varied sources of ‘evidence’ and competing drivers clearly at play (Davies et al., 2008). Sociocognitive perspectives (Landry et al., 2001; Ringberg and Reihlen, 2008) on knowledge transfer alert us to the importance of ‘mental models’ that guide people’s sense-making. There is a need to consider not only technical skills and resources, but also the values, assumptions and worldviews of the actors who generate, and potentially apply, knowledge relating to health inequalities. A series of earlier empirical studies have demonstrated the value of examining the understandings and experiences of these professionals. For instance, research has revealed sharply contrasting epistemological and ideological positions within the health inequalities research community (Garthwaite et al., 2016; Smith and Eltanani, 2015), and the ways in which some health policy makers question whether a consideration of ethnic health inequalities is a legitimate part of their role (Salway et al., 2016). Such investigation can also provide important insight into organisational and institutional contexts; revealing both the explicit priorities, and the more implicit, taken-for-granted modus operandi that shape health inequalities work. Several studies have highlighted the ways in which cultures of evidence, professional hierarchies, and organisational relationships shape (in)action on health inequalities (Lorenc et al., 2014; McGill et al., 2015; Petticrew et al., 2004; Smith, 2014; Turner et al., 2013; Whitehead et al., 2004).

To-date there has been very little exploration of how intersectionality is travelling within health inequalities work beyond North America. In this paper, we contribute to filling this gap by exploring professional stakeholder understandings, perspectives and experiences. We include both researcher and policy/practice stakeholders because bridging the research-implementation gap requires engagement from both sides.

## Methods

A research team comprising university researchers and policy/practice professionals designed and implemented an online survey and a stakeholder workshop.

### Survey

The online survey (Supplementary material) included two questionnaires, one tailored for researchers and one for policy/practice professionals. Shared topics included: familiarity with the term and concept ‘intersectionality’, general reactions to it, and practical issues and barriers to its uptake. Respondents were also asked their opinion on two aforementioned specific policy suggestions – intersectionally targeting and tailoring interventions, and evaluating the intersectional effects of policies. Socio-demographic information was collected. Policy/practice professionals were additionally asked how health inequalities are currently understood within their work arena and were asked questions about a vignette referencing gender, ethnicity, age and socioeconomic background (Supplementary material). Researchers were asked about three specific research challenges – categorisation, intersectional heterogeneity, and policy/practice relevance.

The survey was advertised on academic mailing lists, Twitter, and policy/practice networks. Direct invitations were also sent via existing contacts and respondents were asked to suggest further potential respondents, who were also invited.

### Workshop

The workshop created an opportunity for detailed dialogue around the idea of intersectionality. Specific aims were to: explore the potential of intersectionality for understanding and/or tackling health inequalities; share examples of how intersectionality can be applied in health inequalities practice and research; address challenges; and to identify potential ways to advance intersectional approaches.

We invited people from a range of backgrounds via the survey and direct invitation. The workshop included presentations from practice and research, subgroup discussions, and a concluding plenary discussion. Three note-takers took detailed notes. For the subgroup discussions facilitators were given a topic guide, as well as practical tasks centred on engaging with illustrative survey responses. We aimed for a dialectical engagement at the workshop by feeding survey responses back to participants to expand them in the group setting, allowing for wider reflections.

### Analysis

Open-ended questionnaire responses and notes from the workshop were read multiple times by two researchers to aid familiarisation with the data. A set of initial themes was derived from the survey and workshop topics (in turn based on the literature reviewed above) and expanded inductively from early readings of the data, which were: the term ‘intersectionality’ itself, practical barriers to implementation, and challenges with intersectionality research. We also asked about two specific policy suggestions: (i) targeting and tailoring interventions, and (ii) evaluating the effects of existing policies. The following themes emerged in successive iterations: complexity and consistency, potential for improving patterns and causes of health inequality, operationalising the approach in research and policy analysis, and intersectionality as a prompt for action. Direct quotations from the surveys and segments of workshop notes were coded against these themes.

### Ethics

Ethical approval was obtained from [retracted] for both the survey (reference number 023494) and workshop (reference number 024454) elements of data collection, including informed consent to record data at the workshop.

## Results

### Participant characteristics

53 researchers (95% university-based) and 20 non-researcher stakeholders responded to the survey. For researchers, seniority was varied with responses from PhD students through to full professors. 33 researchers were women and 30 identified as White English/Welsh/Scottish/Northern Irish. Across both the survey and workshop, the policy/practice professionals represented a wide range of roles (e.g. evidence manager, director of communications, equality and diversity manager, administrator, director of public health, health improvement principal) and sectors (e.g. third sector, local government, National Health Service, national statutory agencies). All respondents were based in the UK, with a wide geographical spread, except 4 researchers who were based in Europe. 15 non-researcher stakeholders were women and just under half identified as White English/Welsh/Scottish/Northern Irish. The workshop involved 23 people, and as with the survey involved wide representation in terms of socio-demographic factors and professional backgrounds.

### Current understandings of health inequalities

Policy and practice respondents to the survey were asked to comment on the kinds of explanations people in their field of work give for ‘why some social groups (e.g. according to gender, ethnicity, age, or socioeconomic factors) have better or worse health than others.’ They were then prompted to rank each of the following potential explanations on a scale from 1 (barely or never used) to 5 (dominant explanation): ‘cultural’, ‘behavioural/lifestyle’, ‘political or economic’ and ‘discrimination.’

Responses indicated a range of co-existing explanations within and across work arenas. Explanations rooted in the characteristics or (un)healthy behaviours of individuals and groups were mentioned spontaneously by most respondents. One respondent referred to this as ‘deficit’ language. When prompted, 16/20 thought that behavioural/lifestyle, and 12/20 that ‘cultural’, explanations were dominant (scoring 4 or 5). A particular version of the latter explanation was identified as ‘culture and language’ underpinning ethnic minority health disadvantage.

Most respondents also spontaneously identified ‘social determinants’ or ‘wider determinants’ as commonly articulated explanations for health differences between groups. These were understood primarily in terms of inequalities in material and financial resources, and in some cases place-based deprivation. A couple of respondents noted that inequalities tended to be understood as resulting from behavioural processes, especially in statutory organiastions, with individual behaviours remaining the ‘go-to-target’ for action. A further respondent noted that political dimensions are not often made explicit in such ‘social’ explanations. When prompted, just nine respondents identified ‘political or economic’ factors as dominant explanations.

A further set of explanations mentioned spontaneously related to differential access to health services. Respondents felt that barriers to care, and problems ‘navigating the system’, were commonly employed explanations. In some cases, these understandings were linked to notions of individual obstacles. In others, the understandings appeared to be more structural, with service cut-backs, (in)adequacy and (in)eligibility, being mentioned.

Exclusion and discrimination were less often identified as explanations. One respondent spontaneously referred to ‘minority stress theory’, another to ‘intergenerational adverse experiences’ and another to ‘prejudice and discrimination’, as ways that the causes of health inequalities are understood by some colleagues. When prompted, discrimination was identified as a dominant explanation within their field of work by just two respondents.

After being presented with the vignette (Supplementary material), policy and practice survey respondents were asked to comment on how their area of work deals with these multiple factors, with four possible responses. Intersectionality had not been mentioned in the survey at this point to avoid leading respondents. Eleven respondents answered, ‘it mainly focuses on one attribute (e.g. gender, or ethnicity) at a time’ and a further three answered ‘it mainly focuses on one attribute at a time, but also considers how attributes might be mutually important’. Just six answered; ‘it mainly considers all attributes, but also focuses on one attribute at a time in some cases’ and none that ‘it mainly focuses on all attributes at the same time.’

### Familiarity and appeal of ‘intersectionality’ as a term

The term ‘intersectionality’ was not spontaneously mentioned by any of the practice or policy survey respondents. However, when directly asked whether they were familiar with this term just three reported that they had never heard of it and two that they had only ‘heard a little about it.’ Among the researcher survey respondents, 37/53 reported being ‘fairly’ or ‘very’ familiar with the term, while 12 had heard a bit about it, and four reported that they had never heard of it.

When presented with a definition of the term, 19/20 of the policy/practice survey respondents felt that it ‘made sense’ and had relevance to their work.

However, concerns were expressed across both survey respondent groups that the term itself is off-putting and not “user friendly” nor “plain English.” Around a third of survey respondents in each group felt it was more unhelpful than helpful as a term.

> *“Intersectionality is academic speak and prevents engagement with the public*.” Researcher, survey
>
> *“If someone asked me what I did today I would say it was about looking at ways of tackling inequalities. Calling it intersectionality research may silo it*.*”* Workshop participant
>
> *“I believe that this is a widely understood concept by practitioners in my field, but they would not necessarily adopt the word*.*”* Policy/practitioner, survey

### Conceptual complexity and (in)consistency

In addition to scepticism about the term itself, a dominant theme among survey and workshop respondents related to conceptual complexity. Concerns were expressed regarding a lack of clarity and inconsistency in how intersectionality is understood.

> *“It is a complex idea; it is not necessarily the term that is the problem”* Researcher, survey
>
> *“Intersectionality is an approach with fluid margins*.*”* Researcher, survey
>
> *“I suspect there may be reluctance in applying this concept for several reasons - it is inherently complex*… *the specialist skillset required to use the concept meaningfully is limited*.*”* Policy/practitioner, survey

Furthermore, sharply contrasting responses between policy/practice participants suggested variability in levels of understanding across organisational contexts, with staff in specialist third-sector organisations perhaps having greater knowledge of the concept and its origins than those working in the statutory sector.

> *“I think it is widely understood by practitioners in my team and with partner organisations*.*”* Policy/practitioner, survey
>
> *“A lot of people don’t really understand it, and perhaps dismiss it as post-modern social-justice-warrior work*.*”* Workshop participant

A particular concern expressed by some participants related to maintaining intersectionality’s critical edge; its focus on power, relational dynamics, institutionalised discrimination and systems of oppression, and on transformational change.

> *“Intersectionality is more than simply describing differences in ever more refined disaggregations, but entails an institutional analysis of the occlusion of certain intersections*.*”* Researcher, survey
>
> *“[Intersectionality] has an explicit commitment to social justice that goes beyond just an explanation or description of health inequities but taking that next step towards trying to find change and transformation”* Workshop participant

Comments from other participants tended to confirm the absence of this critical understanding among at least some of those working in research, policy and practice.

> *“Explain to me the difference to “statistical interaction”. Don’t make people learn to speak intersectionality*.*”* Researcher, survey
>
> *“Not sure what added value an intersectionality lens adds to this, as this is how design thinking would approach the problem but would also include capabilities, motivations and opportunities*.*”* Policy/practitioner, survey
>
> *“Feels like a descriptive term as opposed to one that generates action like ‘human rights’*.*”* Policy/practitioner, survey

An area of potential contestation and confusion related to which social identifiers and processes of disadvantage should be in view. For some, intersectionality is about disadvantage associated with minority ethnicity, rather than, say, disability or gender.

> *“Main issue where I work is simply that diversity in our geographic area of focus is very low, therefore even initial discussions around diversity can be difficult, let alone intersectionality*.*”* Policy/practitioner, survey

Other participants were concerned that other axes of difference (notably age and gender) and dynamic processes of disadvantage (including across the life-course) should not be overlooked.

> *“I believe age and generation is an additional factor - periods of greater and lesser equality/ welfare states and their cut backs”* Policy/practitioner, survey
>
> *“I feel you have missed the cumulative aspect of intersectionality. It is a conferred and incremental (dis)advantage*.*”* Researcher, survey

### Potential to improve understanding of patterns and causes of inequality

Most participants felt there was potential for intersectionality to provide new insights regarding patterns of inequality beyond those offered by currently dominant approaches.

> *“An intersectionality approach, would, in my view, enable a better understanding that targeted interventions to address one axis of discrimination, such as gender, may actually make matters worse for certain intra-categorical sub-populations by directing focus and resource away from those in greatest need, whilst giving the illusion of effective action being taken*.*”* Workshop participant
>
> *“We miss high risk groups or inequalities by only considering single statuses or identities*.*”* Researcher, survey
>
> *“Intersectionality therefore offers a way to recognise that multiple factors play an intrinsic role in how individuals interface with their environments, which may offer clues in how to prevent and address health inequalities*.*”* Workshop participant

However, a minority were less convinced of its utility.

> *“We think in a general sense about people who may be disadvantaged. I am not sure that it would be necessary for us to be much more granular, though we are aware that we know little about ethnic minority experiences*.*”* Policy/practitioner, survey
>
> *“I do not think that it really stands in contrast to traditional approaches, it just has a broader focus than traditional ones*.*”* Researcher, survey

Among those participants who considered intersectionality to be an important tool, there was also variation in understandings. For instance, some participants suggested that intersectionality is a framework through which identification of – perhaps previously unrecognised – disadvantage can emerge.

> *“What we would be talking about here is emphatically not university researchers analysing policy outcomes against a list of predefined subgroups. Intersectionality would require community interaction and critical perspectives on what the relevant groups are*.*”* Researcher, survey
>
> *“The whole point of intersectionality is to attempt to recognise complexity - and that the immediately apparent lines of discrimination might not be the only ones that matter*.” Researcher, survey

In contrast, another participant saw the contribution as one of highlighting the circumstances of groups already assumed (or demonstrated) to be severely disadvantaged.

> *“I thought that this questionnaire might be about looking at the really troubling and hard to reach groups impacted upon by health inequalities and am disappointed that it doesn’t (I have worked with older prisoners who are mainly sex offenders and issues of inequality, stigma, shame, discrimination are so sharp for them)*.*”* Researcher, survey

Similarly, mixed opinions were expressed regarding the potential for intersectionality to improve our understanding of the causes of health inequalities. Caution was expressed by some workshop participants that intersectionality may be a new ‘buzzword’ that fails to add value.

However, several participants suggested that an intersectionality approach has the potential for greater attention to processes of ‘group’ formation, that is, how people come to be identified, or to self-identify, with particular social locations and the implications that these have for health. Rather than taking such ‘groups’ for granted, participants felt that intersectionality can help to interrogate their meaning and relevance over space and time, including attention to individual biography and collective histories. Intersectionality was also seen as useful in highlighting agency, and the divergent experiences of people who make up groups labelled as disadvantaged, thereby providing more nuanced understandings.

> *“The impact of the social attributes you have, or establish, may differ by location, affluence and social makeup of the wider community, location and demographics. Generalisability would not be sound*.*”* Researcher, survey
>
> *“We should also not forget (nor overstate) the reflexivity or agency that multiply disadvantaged individuals and groups can deploy”* Researcher, survey

### Operationalising an intersectional approach in research and policy analysis

Survey respondents and workshop participants were also asked to comment on the practicality and feasibility of using an intersectional approach to describe and understand health inequalities, with the majority identifying significant challenges.

Most researchers and policy/practice participants raised the issue of data availability limiting the potential for intersectional approaches to quantitatively describing health inequalities. Large datasets that include multiple social attributes measured well are typically not available. Some intersections are easier to examine than others due to the categories that are typically employed – or overlooked – in routine datasets and research studies. Participants noted that data on ethnicity continues to be of poor quality and completeness in many UK administrative datasets, while data on sexuality, disability and migration status is often totally absent.

Several researcher participants highlighted the specialist statistical expertise needed to undertake technically complex analyses, and the limits of accepted quantitative approaches to health inequalities analyses. This is particularly the case where intersecting variables are highly correlated, making it difficult to conceptually (let alone statistically) separate the effects of those variables

> *“To consider several factors at the same time and not focusing on one can be difficult as the factors correlate with each other and it is difficult to estimate the true effects of single measures*.” Researcher, survey
>
> *“Socioeconomic position and sex or race are not causally independent. The investigation of intersectionality seems to me to be difficult to put in some counterfactual frame in quantitative method*.*”* Researcher, survey

More fundamentally, several survey and workshop participants highlighted perceived dangers of employing quantitative methods alone.

> *“Adoption of the term by quantitative researchers may dilute it through overuse and, due to the often atheoretical (implicit theory) nature of much quantitative research, stripping out of its theoretical grounding and interest in concepts such as marginalisation*.*”* Researcher, survey

Several respondents expressed concern that categorisation may be driven by data availability rather than by prior evidence and associated hypotheses regarding processes of disadvantage impacting upon sub-groups of people.

> *“You would need some evidence to support sub-group choice - so why age 50-55 rather than 50-65 or 40-55 or whatever, why Black women, why low education - need to draw on relevant evidence to support choices*.*”* Researcher, survey
>
> *“Intersectionality would lose its critical edge if it becomes a data-mining exercise in which we search for differences across an infinite number of categorisations*.*”* Researcher, survey

However, a minority of researcher respondents were in favour of exploratory analyses across the range of intersections represented in available datasets.

The importance of moving beyond description towards explanation and modifiable factors was also emphasised across the survey and workshop. Here mixed methods approaches were advocated by many participants for generating understanding of the processes that (re)produce disadvantage.

Limitations of quantitative approaches included difficulties in measuring discrimination operating at structural and institutional levels. Qualitative methods were seen as particularly suitable for grasping the lived experience of intersectional identities and reducing the risk of stigmatisation of marginalised groups by giving them voice and explaining the processes through which health disadvantages come about. Nevertheless, participants felt that effective integration of qualitative and quantitative approaches requires training, support and a change in research culture, with them often remaining in “separate boxes” currently.

> *“Micro-categorisation is a trap. Use a mixed-methods approach. Nothing gives you more heterogeneity than a story, the things that are told and that are not told*.*”* Researcher, survey
>
> *“Let’s not try to explain everything but stick to sufficient and remediable areas of explanation of difference in outcomes*.*”* Researcher, survey

Most researcher respondents agreed that there was value in exploring advantaged positions – the ‘contours of privilege’– as well as marginalised sub-groups.

A number of participants felt that the potential of intersectionality to generate new insights was so far largely untapped. One workshop participant argued that methods lag behind the theory, especially regarding intersectionality’s focus on the relationship between social power and identity, and the need to elucidate dynamic events, contexts and processes. Another felt that academics “shy away from” actually identifying practical solutions to inequalities in processes and outcomes revealed by intersectional analysis.

### A prompt to action

In terms of whether intersectionality can inform more effective action on health inequalities, survey and workshop participants were asked to consider the merits of using intersectionality to target and tailor interventions to the needs and circumstances of specific population subgroups. Opinions were mixed and nuanced, with around two-thirds of survey respondents seeing some merit in such an approach.

Some participants highlighted the important distinction between ascertaining which intersectional categories have the worst outcomes on paper versus identifying population sub-groups that are meaningful and practicable for action in the real world.

> *“There definitely needs to be a move away from one-size-fits-all, so the idea is good in principle. One thing to consider is the extent to which any groups formed actually do share a unique point of view, i*.*e. do members share the same needs in terms of an intervention. For example, in terms of health inequalities policy, is there a meaningful case to [conceptually] isolate those aged 50-55?”* Policy/practitioner, survey

Some noted that targeting by geography, e.g. in specific communities, rather than by other social identifiers, is often more feasible.

A further distinction was drawn between higher level policy on the one hand, and the design and delivery of interventions or services on the other. At the former, more strategic, level tailoring was felt to be more difficult.

> *“Nice in principle, probably impractical in reality. Interventions might be better suited to this but it seems to me policy does not lend itself well to nuance and tailoring*.*”* Researcher, survey
>
> *“Policy makers want things to be kept simple Although I think there is substantial recognition that there are few (if any) cases where a one-size-fits-all solution works, many actors in social policy have yet to figure out how to create policy that effectively accounts for this*.*”* Policy/practitioner, survey

Other concerns with targeted approaches expressed by survey and workshop respondents included: excluding people with needs who do not fall into the targeted category; stigmatising and reinforcing deficit narratives about recipients; fragmented approaches; and addressing downstream factors while leaving upstream inequalities unchanged.

> *“The big issues of race, sex, and class don’t need targeted interventions but structural changes*.*”* Researcher, survey

Some respondents felt that intersectional analysis could generate knowledge to inform effective tailored and targeted responses to need, but only if the approach is operationalised through the use of participatory approaches that effectively include marginalised people.

> *“I think it [the suggestion of using intersectionality to target/tailor interventions] is a good one, especially where it gives more voice, choice and power to people who are less listened too. It is when service providers and policy makers listen to anticipated beneficiaries that they can learn about what can work for them and what are their needs, wants and barriers to achieving good health*.*”* Policy/practitioner, survey

Respondents were more consistently positive about the suggestion that intersectionality could be used to identify and understand the differential effects of existing policies.

> *“I would certainly agree that better work, which incorporates an intersectional lens, should be done to understand the impact of national or local policies on different groups in society. This would contribute to improved policy making*.*”* Policy/practitioner, survey
>
> *“I like it. National and local level policies often disadvantage groups when they are supposedly designed to advantage them. The more these impacts can be articulated the better*.*”* Researcher, survey
>
> *“A more robust understanding of intersectionality, and the methodologies that accompany it, has the potential to better capture the lived experience of people who experience discrimination, at the same time as helping social action have an impact that does not just accumulate benefits to some*.*”* Workshop participant

Again, the importance of community engagement and recognition of power were highlighted by some.

> *“Intersectionality would require community interaction and critical perspectives on what the relevant groups are, the different power dynamics, how and why the policy is having an impact, and whether the outcomes are even the right ones to be measured*.” Researcher, survey

Aside from the generation of knowledge that could theoretically inform better action, several participants questioned whether an intersectionality approach could *influence* the direction and focus of current health inequalities policy.

Suggested obstacles to intersectionality having an influence on policy were a desire for clear and simple solutions, fear of uncertainty, and the predominance of cost-benefit (or even invest-to-save) justifications for action on inequality. The latter was felt to be particularly significant among “cash-strapped public services.”

> *“We struggle to act when we examine one aspect, so this idea may feel overwhelming”* Policy/practitioner, survey
>
> *“An approach/policy devised in this way creates higher budgetary pressures since they reduce the savings from economies of scale. It may be that there needs to be a greater shift from a cost-benefit approach to a rights-based approach”* Policy/practitioner, survey

In addition, some participants emphasised that the prioritisation of socioeconomic inequality over other axes of difference and disadvantage could undermine the perceived relevance of intersectionality to policy-makers. This was linked to organisational structures that lack diversity and cultures that fail to recognise axes of difference and discrimination (across both academia and policy making bodies).

> *“The political context, and probably the lack of diversity in the work place, and the organisational culture, I believe, provide barriers to intersectionality*.*”* Policy/practitioner, survey
>
> *“There needs to be a shift in general because one barrier is that people don’t see how certain factors can lead to inequality. It is as if some kind of ‘inequality blindness’ is built into*
>
> *society*.*”* Policy/practitioner, survey
>
> *“Having a new term in itself does not necessarily move things forward*.*”* Researcher, survey
>
> *“Lack of representation by people who occupy particular social positions and identities. The academy is predominately white middle class which fosters institutionalised racism and classism*.*”* Researcher, survey

And, while some participants offered examples of existing policy work that they felt had incorporated attention to intersectionality, these appeared to be cases where attention had been expanded beyond socioeconomic status as a sole axis, but it was less clear that there had been either a noticeable shift towards a focus on power and processes of marginalisation, or real engagement of those who are marginalised, had been achieved.

> *“My experience is that public health policy often does not engage particularly well with theory or with critical perspectives. It is reasonably willing to engage with the idea of multiple variables (and notions such as dual discrimination under the equality act do allow for this kind of ‘additive approach’). I think it is a lot less willing to engage with questions of epistemology and of power*.*”* Researcher, survey
>
> *“Obstacle isn’t [understanding the concept] - it’s putting emphasis on structural inequalities and social justice rather than an individual approach*.*”* Workshop participant

However, other respondents were more optimistic that intersectionality approaches could be influential in shaping public debates and policies, for instance if they were able to highlight the differential benefits and harms of government action for sub-groups within society. Case studies and real-life narratives were suggested as a way to do this.

> *“Show how it matters. That not all women are the same, not all ethnic minorities and not all lower educated is obvious. But the implications of this fact for public health policies are not and these should be articulated*.*”* Researcher, survey
>
> *“Community activism may be a mechanism for pointing out to policy-makers that there are dynamics and concerns that there are overlooking. However, I have to agree that this is a challenge*.*”* Researcher, survey

## Discussion

Intersectionality has been regarded as a promising framework for understanding and tackling health inequalities. Its explicit acknowledgement of diverse and complex inequalities, and the power structures and systems of discrimination that shape them, are thought to offer the potential to reframe, and prompt more effective action, on health inequalities. However, the academic and policy/practice stakeholders’ perspectives presented above clearly resonate with earlier work, indicating the need for caution, alongside suggestions for ways in which the potential of intersectionality might be realised. The finding that policy and practice stakeholders did not spontaneously mention intersectionality when asked about explanations for health inequalities suggests that it mostly remains a fringe perspective in the UK policy context. Part of this may be due to its centring of complexity – often at odds with the simplicity craved by policy makers – but it may also indicate that it is seen as a more theoretical rather than practical approach, or because it is specifically concerned with issues of equity and social power.

As noted by Hankivksy (2014), a pre-requisite for adopting an intersectionality approach is an openness to social justice, and a willingness to move away from prioritising *a priori* singular axes of inequality. Further, organisational and institutional hierarchies have a strong influence on how health-inequalities policy is developed and implemented (Lorenc et al., 2014; Smith, 2014). Differing perspectives and vested interests mean that different voices in the health inequalities field often engage in ‘boundary work’, questioning the legitimacy of approaches other than their own (Garthwaite et al., 2016). Situating intersectionality within the wider context of health inequalities policy therefore suggests, like any approach, its adoption will be filtered through the existing dynamics of the field. Indeed, the continuing dominance of individualised behavioural explanations was noted by many stakeholders – in many ways anathema to intersectionality’s concern with intersecting systems of discrimination and marginalisation (Brassolotto et al., 2014).

In addition, the contested nature of intersectionality and its ‘theoretical, political, and methodological murkiness’ (Nash, 2008) no doubt contributed to the more divergent and hostile views we observed. Different disciplines and scholars lay claim to what intersectionality is or should be. As noted, some scholars passionately reject analysis of individual intersectional differences divorced from concerns of liberation and justice (Jordan-Zachery, 2007). From such a perspective, analysis of health inequalities according to the ‘big four’ axes of inequality (age, sex, class and ethnicity) is questionable. Some stakeholders regarded intersectionality as an approach of those at the margins i.e. it should focus on typically excluded populations. Related to this is the question of whether focussing on advantaged intersectional positions/identities is warranted, or whether intersectionality should be principally concerned with disadvantage – a clear tension in intersectionality research (Nash, 2008) that was also reflected in stakeholder views. In our view, Bauer (2014:12) offers a sensible solution: consideration of all positions/identities enables researchers to unpack how privilege as well as marginalisation affects health, offering ‘the potential to provide new and interesting observations on the distribution of the burden of disease across social location’. The question arises as to whether the contestation evident among stakeholders and prior literature is inevitable and simply reflects the processes and dynamics that occur with health inequalities research more generally, or whether it is possible that refinement and development of the framework can allow it to ‘travel further’ in the policy and practice world. The development of clearer methodological guidelines and expertise – currently a sticking point in its wider adoption (Bowleg, 2008; Hankivsky et al., 2014) – seems uncontroversial. However, it is arguable that heterogeneity, including in the focus of intersectionality itself, is inherent to the approach given its rooting in standpoint theory and emphasis on multiplicity.

Despite contestation, there was also a general consensus across stakeholders that intersectionality should be concerned with subgroup differences *in relation to* wider interacting systems of social power (what Collins (2002) refers to as the ‘matrix of domination’). This provides a shared foundation upon which methodological work can potentially advance. There is, however, a clear need for the development of better empirical methods that can operationalise intersectionality’s conceptual complexity and successfully explicate mechanisms, processes and contexts. It is unsurprising that many already squeezed policy and practice professionals, as well as researchers, questioned what intersectionality actually adds to existing approaches, and what we might miss by ignoring it. The onus is on researchers to demonstrate the value of the approach and make it accessible for others. This is especially important given the current policy climate where public resources are already constrained. The potential for qualitative and mixed methods work here is substantial, including in demonstrating the value of the approach, capturing complexity and elucidating causal processes. Yet given its potential complexity, it is impossible for each piece of research to cover all bases. For example, advanced quantitative work is unlikely to include a critical participatory perspective, and theoretical work need not take an applied policy approach. In our view, it is justified for specific pieces of work to take a variety of approaches to advance our methodological and empirical understanding, so long as the researchers involved are mindful of intersectionality’s concern with wider systems of discrimination and social power and where possible frame results in these terms. Similarly as Smith (2016:76) notes, ‘no one writer can address all identities directly in a single piece of work, what is needed is recognition of a plurality of voices in mainstream scholarship.’ An efficient division of labour is needed so that different approaches are complementary, held together with the underlying thread of the intersectional paradigm. This is likely to result from training and support and a move towards commonality in concepts and language.

With respect to quantitative research in particular, stakeholders clearly articulated the need for good quality large-scale data. The recent growth of ‘big data’ e.g. biobanks and linked administrative data may provide the numbers needed for highly granular analysis, but this risks being divisive in emphasising fine-grained differences rather than commonalities (the ‘infinite regress’ (Davis, 2008) categorisation problem, especially when those differences are not particularly conceptually meaningful). In addition, such datasets often contain poor measures of anything other than basic social categories, with sexuality for example often absent. In relation to ethnicity, categories and labels need to be meaningful in terms of the specific research questions being explored (Mir et al., 2013), and data on ethnicity are frequently missing or of poor quality. Furthermore, such large-scale datasets rarely include variables that allow us to go beyond describing patterns of intersectional inequalities to unearthing mechanisms, making it difficult to expose and address power and discrimination. With respect to quantitative analysis, we observed the conflation between intersectionality and the use of interaction effects previously highlighted by Bauer and Scheim (2019). While it is necessary to consider statistical interaction in intersectionality research, it is not sufficient, because as noted intersectionality is not the testable hypothesis that there are multiplicative interactions between socio-demographic factors but rather it is a framework, perspective, or paradigm (Bowleg, 2012) concerned with the relationship between subgroup heterogeneity and social power, which could exist with only a combination of additive effects (Holman et al., 2020). This confusion reinforces the need for developing training and expertise in the use of quantitative intersectionality.

How might intersectionality be practically implemented in policy approaches? We explored two potential approaches with stakeholders: intersectional targeting and tailoring of public health policies/interventions, and evaluating intersectional effects of pre-existing policies. Whilst stakeholders thought that the former sounded good in principle and there is a need to move beyond one size fits all approaches, there was uncertainty whether intersectionality can help in doing so. Geographical targeting of interventions was felt to be an effective way to target action (though this raises the question of other characteristics cutting across localities), and participatory approaches thought essential given intersectionality’s concern with marginalised populations. Stakeholders recognised that there are many potential pitfalls and unanswered questions regarding the first approach, around unexplained heterogeneity, stigmatisation, and ignoring the capacity for personal agency and reflexivity. For example, Affirmative Action Plans in the United States, which are differentiated by axes of inequality, have been criticised for the stigmatising effects resulting from perceptions of low self-competence and perceived stereotyping by others (Leslie et al., 2013). Stakeholders suggested that qualitative and participatory approaches (e.g. case studies and real-life narratives) may help mitigate these risks by co-producing responses that appropriately meet need. We found intersectionally evaluating pre-existing policies was seen much more favourably, though this might (rightly or wrongly) lead to the conclusion that targeting/tailoring is needed to avoid such differential impacts. Reactions to targeting/tailoring can reveal implicit understandings about entitlement and (un)deservingness, and what is considered justifiable action to meet the needs of people who are ‘different’ from the White, able-bodied, heterosexual etc. majority. Either way, intersectional approaches to policy evaluation would highlight the danger of attempting to find a single, homogeneous effect of policy, as might be found by standard econometric policy evaluation techniques, rather than understanding that such effects are often different for different sorts of people.

Complexity is perhaps both intersectionality’s biggest asset and challenge. A key question is how to work with the ‘right’ level of complexity, to acknowledge complex social reality and associated inequalities, whilst not being so complex that clear and direct policy and action is inhibited. This is a tricky balance to strike because the level of complexity that allows understanding of inequalities and their causes may not be the same level as that which can be used to formulate and get political backing for a particular policy response. Numerous stakeholders re-iterated the simplicity and efficiency preferred by policy makers. Such viewpoints accord with the idea that in some cases engaging in ‘strategic essentialism’ to stress commonality - for instance according to gender - can help to mobilise action (Smith, 2016). In public health, the concept of proportionate universalism raised by one participant suggests a way in which the targeted might be combined with the universal, to balance the simplicity that policy makers desire alongside a need to consider diversity, differential need and inclusion. Marmot et al. (2010) coined the phrase proportionate universalism to advocate for universal policy action that is also targeted proportionate to the level of need. A strength of this approach in the UK context is that the NHS is founded on the same principle and has wide public acceptance and appeal. Carey et al. (2015:4) outline a framework for how this might be achieved, arguing that ‘an appropriate balance can be struck which guarantees principles of equality and fairness (central to the social gradient approach), with the need to allow for diversity and difference (i.e. effective targeting for different social groups)’.

The research community must, therefore, make intersectionality research accessible if the approach is to be further promoted. One way forward may be to produce a tool or set of guidelines for research and policy audiences on how they can incorporate intersectionality into their work. Hankivsky’s (2014) framework represents a significant step forward, though further developments might pay more attention to empirical and methodological issues – especially in a quantitative framework – and their policy relevance. As noted, a limiting factor may be that intersectionality requires expertise in varied dimensions of health inequality. For example, Marmot barely addresses ethnic health inequalities despite highlighting their importance (Salway et al., 2010). In ongoing work, the authors are contributing to a revision of the Health Inequalities Assessment Toolkit (Porroche-Escudero and Popay, 2020) that aims to support health researchers to incorporate an intersectional perspective into their work.

Alongside developing new guidelines and toolkits, workshop stakeholders suggested integrating intersectionality into wider existing policy/action frameworks. At present, current frameworks tend to acknowledge multiple types of discrimination but only in isolation. For example, Article 13 of the EU Treaty of Amsterdam ‘necessitates that member states must protect citizens from discrimination on a number of grounds including gender, race or ethnic origin, religion or belief, disability, age, and sexual orientation’ (Hankivsky 2011). Similarly the UK Equality Act only allows discrimination claims on the basis of single characteristics. A recent attempt outlining an intersectional approach to human rights law from de Beco (2017) offers a step in the right direction. As Smith (2016) notes, the Joint Equality and Human Rights Forum (JEHRF) and the Advisory, Conciliation and Arbitration Service (ACAS) have examined how intersectional identities affect experiences, suggesting multiple discrimination is common. The suggestion of integrating intersectionality into the WHO Health Impact Assessment is another potential way forward. This will require mobilisation across disciplines and sectors, in recognition of the breadth of the determinants of health.

## Conclusion

Intersectionality has become a buzzword in recent years, regarded as holding great potential to advance research and policy action on inequality. This interest has now clearly landed in the health inequalities field. It is important to sense-check claims regarding the framework by those who work in the field to get a sense of whether these claims match their perspective. We found that, whilst many researchers and policy/practice professionals see the potential value and importance of an intersectional perspective, not all are positive. Numerous obstacles and challenges with the approach were raised, reflecting its relative newness as applied to health inequalities. The views of policy/practice professionals suggest intersectionality has far to travel to help counter individualistic narratives and encourage an approach that is sensitive to subgroup inequalities and the processes that generate them. An appetite for an approach rooted in social justice is necessary. The ongoing COVID-19 pandemic may give new impetus to intersectionality by exposing the scale of unequal impact by ethnicity, deprivation, gender, and age, and prompting debate as to these factors overlap and interact (The Health Foundation, 2020). The public and policy imagination is now surely a more fertile ground for an intersectional approach. Examples of promising practice, albeit mostly in North America, suggest that it is possible for intersectionality to gain traction. The price of intersectional-blindness is potentially significant; it carries the risk that the injustices faced by particular subgroups is missed.

## Supporting information

Researcher survey

Non-researcher survey

## Data Availability

Data is disclosive and has not been anonymised; ethical agreement does not include the public sharing of data.

## Acknowledgements

This work was supported by the Economic and Social Research Council [Grant No. ES/R00921X/1]. Sarah Salway’s contribution to this project was in part supported by the National Institute for Health Research (NIHR) School for Public Health Research [Grant No. PD-SPH-2015-10025]. The views expressed are those of the authors and not necessarily those of the ESRC, NIHR or the Department of Health and Social Care. We are grateful to survey respondents and workshop participants for their insightful thoughts and ideas. We also thank Dr Lois Orton and Dr Calum Webb for their helpful feedback on earlier versions of this paper.

