## Supplementary material for "Can intersectionality help with understanding and tackling health inequalities? Perspectives of professional stakeholders": Researcher survey

### Social groups and health inequalities survey

Welcome to the 'Social groups and health inequalities survey'

This survey is being conducted to understand what those involved in health inequalities research think about how we might understand and explain social group differences in health.  We will explain more on what we mean by this during the survey.

We hope you will take a moment to complete the survey (it should take around 15 minutes). Your participation is voluntary and your response is anonymous unless you decide to share your email address because you are interested in further correspondence.  In all cases, your response will be kept confidential. If you have any questions or comments about the survey please feel free to contact Dr. Daniel Holman, the study's Principal Investigator.

This survey has received ethics approval from The University of Sheffield.  By completing the survey, you consent to having your responses recorded, stored and used for research purposes.  You are free to quit the survey at any time.

Please answer the questions as openly and honestly as possible.  We are very grateful for your time and help.

Dr. Daniel Holman, Professor Sarah Salway, and Dr. Andy Bell (The University of Sheffield)

#### About the concept

How familiar are you with the term ‘intersectionality’?

- ◯ Never heard of it
- ◯ Heard a little about it, but do not really know much about the concept
- ◯ Fairly familiar with the concept; know the basics around what the concept describes
- ◯ Very familiar with the concept; could explain some of the history/background/arguments etc.

Please read the following short description carefully as it represents our basic understanding of the term intersectionality in the context of health inequalities, and will be the basis for further questions

Intersectionality addresses the fact that each of us has a particular gender, ethnicity, age and socioeconomic position. These 'social attributes' overlap and interact with each other to give us a particular *position* in the social structure, and a particular *identity*, shaping our sense of who we are. These positions and identities influence the types of inequalities we might experience. Intersectionality scholars are particularly interested in discrimination and marginalisation, such as sexism, racism and classism, which themselves work together to shape the life chances of those who are for example, female, black and from a disadvantaged socioeconomic background. This understanding stands in contrast to traditional approaches to health inequalities which have tended to focus on one attribute at a time such as ethnicity or socioeconomic position.

- ☐ Please tick the box to confirm you have read the description

What is your initial reaction to our description? If you were already familiar with the term, is it a fair representation? Is there anything you would change or add? If you were not already familiar with the term, does this description make sense?

Your answer

Are there any practical issues or barriers with respect to intersectionality becoming a more widely used concept in research?

Your answer

To what extent do you like the term 'intersectionality' itself in how it describes how social attributes like gender, age, ethnicity together shape people's health?

| | 1 | 2 | 3 | 4 | 5 | 6 | 7 | 8 | 9 | 10 | |
|---|---|---|---|---|---|---|---|---|---|---|---|
| The term is awkward, confusing, or otherwise unhelpful | ○ | ○ | ○ | ○ | ○ | ○ | ○ | ○ | ○ | ○ | It is ideal in what it is trying to describe |

If you think other existing words or concepts would be preferable, please list them here

Your answer

#### Challenges with intersectionality

There are a number of challenges with applying intersectionality to research. We would like to get your views on these to see what the way forward might be. Here we outline three key challenges and then ask for your reaction on each:

##### Challenge 1 - Infinite categorisations

In theory, if you add enough categorisations, gender, ethnicity, socioeconomic position, age, but also disability, nationality, sexuality, location, religion... etc you end up with infinite categorisations such that 'each person is different'. How do we sensibly define intersectional positions? What should we be focusing on? Should we be interested in advantaged as well as disadvantaged positions/identities?

Your answer

#### Challenge 2 - Unobserved heterogeneity

Even within a particular intersectional identity, say 50-55 year old white males with low levels of education, there will be significant heterogeneity in terms of a particular outcome - say rates of diabetes. Is the solution to add further categorisations to account for this heterogeneity, producing ever finer distinctions, or is accounting for this heterogeneity in fact a red herring, and the real strength of intersectionality theory lies elsewhere?

Your answer

#### Challenge 3 - Policy and practice relevance

Intersectionality might sound like jargon to some; the theory can be a little abstract, and the methods obtuse. What is the way forward in encouraging those working in for example public health policy, to consider that e.g. not all women are the same, ethnic minorities are differentiated according to socioeconomic factors, that ageing is an unequal process? Note, it is perfectly fine if you think intersectionality does not have value here, but if so please give some rationale.

Your answer

### Current suggestions on how intersectionality can help with health inequalities policy

Various scholars have suggested intersectionality represents a potential way forward with thinking about policy efforts around health inequalities. We would like to get your opinion on two key suggestions.

One suggestion is that intersectional groups, e.g. 50-55 year old ethnic minority woman with low education, can be used as a starting point to design interventions or policies that are targeted and tailored for specific subgroups. This would move us from a 'one size fits all' to a 'what works for whom' approach. What do you think to this idea?

Your answer

Another related suggestion is that intersectionality can be used to evaluate the effects of national or local level policies on particular subgroups e.g. who is most advantaged or disadvantaged by these policies. What do you think to this idea?

Your answer

#### Next steps

Would you be interested in attending a workshop about intersectionality? The focus would be on similar issues to those asked about in this survey, with the aim to bring researchers and policy/practice audiences together to discuss the concept and whether it can do useful work to tackle health inequalities. If so, please enter your email address below. Note that indicating you are interested does not commit you at this stage.

Your answer

Are you interested in receiving updates on the project findings? If so, please enter your email address below if you did not do this for the previous question, otherwise just enter 'yes'

Your answer

Do you know anyone else who might be interested in completing this survey? If so, please enter their name(s) & position(s) or e-mail address(es) here

Your answer

#### Anything else?

Is there anything else we haven't asked or anything else you would like to share about this topic?

Your answer

#### About you

Are you male or female?

- ◯ Female
- ◯ Male
- ◯ Prefer not to say
- ◯ Other:

What is your ethnic background?

Choose

If you selected any of the 'other' options above, please enter details

Your answer

Which of the following best describes your role?

- ◯ Professor
- ◯ Senior lecturer, assistant professor or reader
- ◯ Lecturer
- ◯ Senior postdoctoral researcher
- ◯ Postdoctoral researcher
- ◯ PhD student
- ◯ Researcher not in academia

How many years have you been working in research? (including PhD if appropriate)

Your answer

Get link

Never submit passwords through Google Forms.

This form was created inside University of Sheffield. Report Abuse

Google Forms
