## Supplementary material for "Can intersectionality help with understanding and tackling health inequalities? Perspectives of professional stakeholders": Non-researcher survey

### Social groups and health inequalities survey

Welcome to the 'Social groups and health inequalities survey'

This survey is being conducted to understand what those involved in policy and practice in relation to health inequalities think about how we might understand and explain social group differences in health. We will explain more on what we mean by this during the survey.

We hope you will take a moment to complete the survey (it should take around 15 minutes). Your participation is voluntary and your response is anonymous unless you decide to share your email address because you are interested in further correspondence. In all cases, your response will be kept confidential. If you have any questions or comments about the survey please feel free to contact Dr. Daniel Holman, the study's Principal Investigator.

This survey has received ethics approval from The University of Sheffield. By completing the survey, you consent to having your responses recorded, stored and used for research purposes. You are free to quit the survey at any time.

Please answer the questions as openly and honestly as possible. We are very grateful for your time and help.

Dr. Daniel Holman, Professor Sarah Salway, and Dr. Andy Bell (The University of Sheffield)

#### Health inequalities amongst social groups

We would like to understand your experiences of how people in your area of work typically understand and explain health inequalities amongst social groups

In relation to your field or type or work, what kind of explanations do people give for why some social groups (e.g. according to gender, ethnicity, age, or socioeconomic factors) have better or worse health than others? Please try to give as much detail as possible

Your answer

To what extent do you think cultural explanations are given? (e.g. norms/values/beliefs/upbringing)

| | 1 | 2 | 3 | 4 | 5 | |
|---|---|---|---|---|---|---|
| Barely or never used | ○ | ○ | ○ | ○ | ○ | Dominant explanation |

To what extent do you think political or economic explanations are given? (e.g. opportunities are unequal, society is unfair, people are exploited by governments or corporations)

| | 1 | 2 | 3 | 4 | 5 | |
|---|---|---|---|---|---|---|
| Barely or never used | ○ | ○ | ○ | ○ | ○ | Dominant explanation |

To what extent do you think behavioural/lifestyle explanations are given? (e.g. diet, alcohol, smoking, exercise, sleep)

| | 1 | 2 | 3 | 4 | 5 | |
|---|---|---|---|---|---|---|
| Barely or never used | ○ | ○ | ○ | ○ | ○ | Dominant explanation |

To what extent do you think reasons relating to discrimination are given? (e.g. sexism, racism, ageism, classism)

| | 1 | 2 | 3 | 4 | 5 | |
|---|---|---|---|---|---|---|
| Barely or never used | ○ | ○ | ○ | ○ | ○ | Dominant explanation |

#### A (very) brief life story: Maya

Please read the following short description carefully as it will be the basis for further questions

Maya is a 58-year-old Indian woman who was born in Lancashire just after her parents migrated to the UK from India in the 1960s. Job opportunities were limited at the time, apart from in local textile industries which were booming. Her mother went to work in a local clothing factory, and her father managed to find work as a clerk at the post office. Maya did not enjoy school much, and part of the reason was that she was called racist names as there weren't many Indian school children in those days. At the age of 16, wanting to earn some money, Maya left to work in the local clothing factory, following in her mothers' footsteps. Maya's wages only afforded her a modest lifestyle, though she managed to buy a small house with her partner. They could afford the necessities, but not much else. She is not looking forward to retirement as she will have to rely on the state pension, though her husband managed to build up a moderate pension working in the post office.

☐ Please tick the box to confirm you have read the description

The story describes Maya's gender, ethnicity, age and socioeconomic background. Some researchers would suggest that these are all important in explaining Maya's health and any illness she might experience. Others would argue that some factors are more important than others, so it is OK to focus one at a time. What is your opinion on this?

- ◯ We should focus on one attribute (e.g. gender, or ethnicity) at a time
- ◯ We should focus on one attribute at a time, but also consider how attributes might be mutually important
- ◯ We should consider all attributes, but also focus on one attribute at a time in some cases
- ◯ We should focus on all attributes at the same time

What is your opinion on what actually currently happens in your area of work?

- ◯ It mainly focuses on one attribute (e.g. gender, or ethnicity) at a time
- ◯ It mainly focuses on one attribute at a time, but also consider how attributes might be mutually important
- ◯ It mainly considers all attributes, but also focuses on one attribute at a time in some cases
- ◯ It mainly focuses on all attributes at the same time

#### Intersectionality

One proposed concept to understand how health inequalities might arise from people having multiple 'attributes' (gender, ethnicity, age, socioeconomic background) at the same time is 'intersectionality'.

Are you familiar are you with the term ‘intersectionality’?

- ○ Never heard of it
- ○ Heard a little about it, but do not really know much about the concept
- ○ Fairly familiar with the concept; know the basics around what the concept describes
- ○ Very familiar with the concept; could explain some of the history/background/arguments etc.

Please read the following short description carefully as it represents our understanding of the term intersectionality in the context of health inequalities, and will be the basis for further questions

Intersectionality addresses the fact that each of us has a particular gender, ethnicity, age and socioeconomic position. These 'social attributes' interact with each other to influence the types of inequalities we might experience. Intersectionality scholars are particularly interested in types of discrimination and marginalisation, such as sexism, racism and classism, which themselves work together to shape the life chances of those who are for example, female, black and from a disadvantaged socioeconomic background. This understanding stands in contrast to traditional approaches to health inequalities which have tended to focus on one attribute at a time such as ethnicity or socioeconomic position.

- ☐ Please tick the box to confirm you have read the description

What is your initial reaction to what we have described; does it make sense to your type of work e.g. in terms of your day-to-day activities, or more broadly in terms of the overall aims/nature of your role?

Your answer

#### About you

Finally, it would really useful to understand a bit about your background

Are you male or female?

- ○ Female
- ○ Male
- ○ Prefer not to say
- ○ Other: ______

What is your ethnic background?

Choose ▾

If you selected any of the 'other' options above, please enter details

Your answer

What is your role?

Your answer

In what type of organisation or sector do you work? E.g. third sector, public health policy, clinical practice, local council, policy making

Your answer

How many years have you been doing this type of work?

Your answer

Get link

Never submit passwords through Google Forms.

This form was created inside University of Sheffield. Report Abuse

Google Forms
